# Analysis of Post-traumatic Stress Disorder Gene Expression Profiles in a Prospective, Community-based Cohort

**DOI:** 10.1101/2023.04.08.23288309

**Authors:** Jan Dahrendorff, Agaz Wani, Thomas E. Keller, Don Armstrong, Annie Qu, Derek E. Wildman, M. Carmen Valero, Karestan C. Koenen, Allison E. Aiello, Monica Uddin

## Abstract

Post-traumatic stress disorder (PTSD) is a common and debilitating psychiatric disorder that may occur in individuals exposed to traumatic events such as accidents, interpersonal violence, war, combat, or natural disasters. Additionally, PTSD has been implicated in the development of a variety of chronic conditions including cardiovascular and metabolic diseases, suggesting that the biological alterations of the disorder can manifest themselves as chronic diseases in those suffering from PTSD. The biological underpinnings of the disorder are not well understood. Gene expression studies can illuminate the complex physiology of PTSD reflecting the embodiment of trauma, i.e. the process in which traumatic experiences in our social environments are manifested in our body by genomic mechanisms. To date, gene expression studies that examine the whole transcriptome are scarce and limited to single-timepoint assessments. Here we applied a transcriptome-wide gene expression screen with RNA-sequencing to whole blood samples to elucidate the gene expression signatures associated with the development of PTSD. The study participants (N=72, of whom 21 eventually developed PTSD) are a trauma exposed subsample of participants enrolled in a longitudinal and prospective cohort study of adults living in Detroit, Michigan. PTSD was assessed in a structured telephone interview and whole blood samples were taken both before and after trauma exposure. We found 45 differentially expressed genes associated with PTSD development with an estimated log_2_ fold change > 1.5 at a nominal p value (p<0.05), however, none of these survived correction for multiple hypothesis testing. Six of the 37 upregulated genes including *PAX6*, *TSPAN7*, *PXDN*, *VWC2*, *SULF1* and *NFATC4* were also ubiquitously expressed in all brain regions examined. Subsequent gene set enrichment analysis identified several pathways relating to brain and immune functioning to be enriched in individuals developing PTSD. Longitudinal sampling provides a promising mean to elucidate the pathophysiology underlying the embodiment of trauma.

---

Post-traumatic stress disorder (PTSD) is a pervasive and debilitating mental disorder with a lifetime prevalence of approximately 6.8-7.3% in the United States general population (Roberts et al., 2011; Kessler & Wang, 2008), while a higher rate on the order of 25-30% has been reported in segments of the population exposed to persistent psychological stress including combat exposed veterans, victims of assault or refugees (Breslau et al., 1998; Kessler & Wang, 2005; Roberts et al., 2011; McLaughlin et al., 2015; Fulton et al.,2015; National Institute of Mental Health, 2017). The disorder manifests itself through a range of symptoms including the recurrent and intrusive recollection of the traumatic experience, changes in arousal and reactivity, alterations in mood and cognition, avoidance of stimuli associated with traumatic event and impairment of functioning in social and occupational contexts (American Psychiatric Association, 2013).

PTSD can have a significant impact on an individual’s quality of life caused by the condition itself or other medical comorbidities associated with the disorder including chronic inflammation (Speer et al., 2018), cardiovascular disease (Edmondson and Känel, 2017), metabolic disease (Talbot et al., 2015) and syndromes (Solomon et al., 2017 and Heppner et al., 2009), substance abuse and suicide (Kessler et al., 2005). Several of these medical comorbidities are exacerbated by a range of poor health behaviors observed in individuals with PTSD including lack of exercise, unhealthy diets, increased tobacco and alcohol use leading to poor metabolic and cardiovascular outcomes (van den Berk-Clark et al., 2018). Recent literature has identified several different pathways in which the condition invokes biological alterations that manifest themselves in cardiovascular and metabolic outcomes including stress regulation and related physical abnormalities reflecting the embodiment of trauma (van den Berk-Clark et al., 2018).

A range of risk factors for PTSD has been previously identified including the level of previous stress exposures, low socio-economic status, extent of social support and female sex (Yehuda & LeDoux, 2007; Brewin et al.,2000; Lowe et al., 2014). The exposure to a traumatic event is the etiological component of PTSD and exposure to such traumatic events is ubiquitous in societies around the world, although it is not deterministic as not everyone experiencing trauma develops PTSD (Resnick et al., 1993; Kessler et al. 1995, Mills et al., 2011; Kessler et al.,2017). In fact, only a small proportion of individuals exposed to traumatic events develop PTSD while the majority remains resilient sometimes even after repeated exposure (Zohar et al., 2009). Recent assessments of the World health organization’s world mental health surveys spanning over 24 countries with 68,894 respondents suggest 70.4 % have experienced at least one lifetime trauma with the most frequently experienced types of trauma in the United States including interpersonal violence and sexual violence in women, while accidents or combat exposures are the most prevalent exposures in men (Benjet et al., 2016). On a global scale, natural disasters, wars and other humanitarian emergencies account for the majority of traumatic experiences (Kessler et al., 2017).

The significant individual differences in response to trauma and subsequent PTSD might be explained by the heterogeneity at the molecular level that can impact someone’s response to trauma. Consequently, identifying the molecular underpinnings influencing psychiatric risk and resilience factors in PTSD has emerged as a priority in the research of the disorder. An enhanced understanding of the complex mechanisms underlying a maladaptive trauma response and the subsequent development of PTSD can help to mitigate the vast morbidity and burden associated with the disorder and might also inform efforts of treatment development, interventions and preventive strategies. Similarly, exploring the potential mechanisms of how lived traumatic experiences are embodied into our physiology and therefore how trauma gets under the skin, can grant insights into how social adversity gives rise to psychopathology. From an epidemiological perspective embodiment can be characterized as a process in which we, as living organisms, biologically incorporate our social and ecological environment (Krieger 2005). Over the last decade significant progress has been made in characterizing the etiologic factors of PTSD. A large body of literature spanning across twin studies, targeted candidate gene analysis, genome and epigenome-wide association studies, as well as methylation and gene expression studies have implicated the role of genetics and epigenetics in the vulnerability to the development of PTSD (Logue et al., 2013; Mehta et al., 2013; Sarapas et al., 2011; Xie et al., 2013). Several risk loci associated with PTSD have been identified (Nievergelt et al., 2019; Smith et al. 2020), however, discrete diagnostic biomarkers underlying PTSD etiology and the pathways leading to the disorder remain to be elucidated.

Emerging literature acknowledges altered gene expression as an important genomic factor in the susceptibility to PTSD (Mehta et al., 2020). The assessment of gene expression can provide valuable insights into the complex pathophysiology of psychiatric disorders like PTSD. However, to date the vast majority of studies has been cross-sectional, preventing identification of pathways and related loci that are involved with PTSD onset rather than a consequence of PTSD. One notable exception to date is a study by Breen and colleagues, who investigated the transcriptome wide gene expression RNA-seq data in peripheral blood of 94 male U.S Marines pre- and post-deployment constructing co-expression networks associated with PTSD (Breen et al., 2015). Their analysis identified modules with an overexpression of genes that were enriched for processes involved with innate immunity and interferon signaling to be associated with PTSD development (Breen et al., 2015). To our knowledge, prospective longitudinal designs as utilized by Breen and colleagues have yet to be applied to a community-dwelling cohort of individuals developing PTSD. Application of these longitudinal study designs would be especially informative, as it is unclear whether gene expression patterns are reflective of current symptoms or indicative of enduring susceptibility prior to trauma exposure. The assessment of gene expression patterns longitudinally is therefore important for the characterization of the molecular underpinnings of PTSD and the discovery of biomarkers with clinical utility.

This study aims to interrogate the gene expression profiles of individuals with PTSD in a community-dwelling cohort. The analysis is based on exploring the transcriptomic profiles in whole blood of PTSD diagnoses based on two different variables focused on PTSD and also a PTS symptom severity construct (explained in the methods below) potentially capable of a more nuanced distinction between PTSD cases and controls. Of particular interest is the investigation of differential gene expression signatures associated with PTSD development, and thereby embodiment of trauma.

## Materials and Methods

### Study Participants and Blood Sampling

The study participants were enrolled in a five yearlong study called the Detroit Neighborhood Health Study (DNHS). The DNHS is a prospective and longitudinal cohort study of adults residing in Detroit, Michigan (Uddin et al., 2010). The initial aim of the study was to evaluate to what extent ecologic stressors including concentrated disadvantage, distribution of income and residential segregation contribute to PTSD risk. Data collection for the DNHS began in 2008 with each wave of measurement including a phone interview over 40 minutes. The participants were compensated for their participation with $25 after each wave. Biospecimen collection was compensated with an additional $25. Data collection for the baseline wave occurred 2009-2010 with approximately annual follow-ups until 2014. At the beginning of each interview and when a biological specimen was collected informed consent was obtained. The study protocol and the DNHS has been reviewed and approved by the Institutional Review Board of the University of Michigan and the University of North Carolina-Chapel Hill. A subset of DNHS participants consented to provide a biospecimen. The biospecimens were acquired from the participants during an in-home visit by a phlebotomist. The samples were subsequently shipped and processed at Wayne State University in Michigan (Uddin et al., 2010). From this biospecimen subset 72 trauma-exposed participants, out of which 21 developed PTSD in the follow-up wave, were identified along with 144 available whole blood samples taken both before and after trauma exposure (e.g from wave 2 and wave 4 respectively). Of the 72 study participants, 70 experienced at least one new trauma between the baseline and the follow-up measurement. The two participants who were not exposed to a new trauma between measurements were among 69 participants who experienced at least one trauma prior to the baseline measurement, such that all 72 participants were trauma exposed by the follow-up wave (wave 4). Total RNA was isolated from the leukocytes of the blood samples with Leukolock kits (Ambion, Austin, TX), as previously described (Logue et al., 2013).

### Assessment of PTSD and Demographics

PTSD symptoms were assessed using a modified version of the PTSD Checklist (PCL-C) via a structured telephone interview (Weathers & Ford, 1996) validated against the CAPS (Uddin et al., 2010). The PCL-C is a 17-item measurement of PTSD symptoms that uses the PTSD symptom criteria from the Diagnostic and Statistical Manual of Mental Disorders (DSM-IV) (American Psychological Association [APA], 1994). The telephone interview also included a range of additional questions regarding the duration, timing and impairment of the symptoms to identify probable cases of PTSD according to DSM-IV criteria. Additionally, a list of specific traumatic events was presented from which the participants were asked which of them were experienced in the past (Breslau et al., 1998). If all six DSM-IV criteria were met by participants at some point in their lifetime, they were considered affected by lifetime PTSD. In addition to the assessment of PTSD, demographic variables including race, sex and age were collected.

Additionally, a post-traumatic stress (PTS) symptom severity measure was also explored as a measure of traumatic stress that could potentially identify gene expression signatures regarding PTSD development in individuals who have experienced PTS but not to an extent consistent with a PTSD diagnosis according to DSM-IV criteria. An increase of statistical power is achieved because PTS symptom severity is a continuous variable while PTSD is a dichotomous variable based on the fulfillment of all PTSD diagnostic criteria.

### RNA-Sequencing

RNA-sequencing (RNA-seq) was performed on all samples (n=144). The ribosomal RNAs were removed with Ribozero Human/Mouse rat from Illumina. The RNAseq libraries were prepared with TruSeq stranded total RNA-sequencing Sample Preparation kit by Illumina. The libraries were quantified by qPCR and subsequently sequenced on 8 lanes for 101 cycles from each end of the fragments on a HiSeq 4000 sequencing system (Illumina) along with the HiSeq 4000 sequencing kit version 1. Each of the samples had a least 60 million reads. Fastq data was generated and demultiplexed with the bcl2fastq v2.17.1.14 Conversion Software (Illumina). For bioinformatic analysis, the resulting data was quality controlled using FastQC (Andrews, 2010), version 0.11.9. Sequencing adaptors have been trimmed from the reads. The resulting reads were aligned to the reference genome GRCh38 (ftp://ftp.ebi.ac.uk/pub/databases/gencode/Gencode_human/release_24/GRCh38.primary_assembly.genome.fa.gz). After alignment of the reads the quantification of expression on the gene level was facilitated with the use of the Featurecounts function of the R-package Subreads, version 2.0.0 (Liao, Smyth & Shi, 2019) on R version 4.0.5. (03, 2021) using Bioconductor version 3.15. The resulting read count data from the Featurecounts table was filtered by the number of counts and the low read counts were subjected to removal prior to downstream analysis to exclude genes with significantly low gene counts across all libraries as they provide insufficient evidence for differential expression. Genes were filtered on the basis of counts-per-million (CPM) with the use of the R-package edgeR, version 3.15 (Robinson, McCarthy & Smyth, 2010), retaining genes if they are expressed at CPM above 0.5 in our samples. Retaining genes expressed at this particular threshold has been recommended in previous literature of PTSD and the authors of the edgeR program (Kuan et al., 2017; Robinson, McCarthy & Smyth, 2010). After the filtering step, edgeR was used to calculate the normalization factors used to scale between the libraries (Robinson, McCarthy & Smyth, 2010).

### Blood Cell-Type Deconvolution and Transformation of Cell Proportions

Differences in cell type proportions and composition as well as their contributions to gene expression profiles is an important determinant of phenotypic variation capable of influencing disease states (Sharma et al., 2019). The identification of discrete cell populations is also important in analysis of differential gene expression as it can be substantially confounded by differences in the cell proportions in each sample. To address this issue, in this study the deconvolution of various blood cell types was performed with Cibersortx, a software applying a computational framework based on support vector machine regression to infer different cell types and cell type specific expression profiles in bulk RNA samples without relying on physical cell isolation (Newman et al., 2019). To enumerate the fractions of various cell populations in the RNA-sequencing data with Cibersortx two input files were required. The first required input is termed “signature matrix” a knowledgebase representing marker genes enabling the discrimination between different cell types and subsets. Further, a tab-delimited text file called “mixture file” was utilized consisting of the gene expression profile counts of the samples.

The reference signature matrix “Non-small cell lung cancer PBMCs” is based on single-cell RNA-seq analysis derived from peripheral blood mononuclear cells previously isolated from tissues of patients suffering from non-small cell lung cancer generated on the 10x Genomics Chromium v2 assay (Chen et al., 2020). The matrix consists of profiles of six leukocyte subsets including B Cells, CD4T & CD8T Cells, monocytes, natural killer cells and natural killer T cells. This reference signature matrix has been previously used in another RNA-seq study investigating multisystem inflammatory syndrome in children (Beckmann et al.,2020).

Given a computational approach was used to deconvolute the cell fractions of the five cell types, the issue that our five cell type fractions add up to 1.0 for the samples has the implication that the cell fractions are only spanning four dimensions precluding knowing how four fractions determines the fifth (Hoffman et al., 2022).This issue was addressed by transforming the five cell fractions into four variables with the use of the isometric log ratio of the R package compositions (Van den Boogaart & Tolosana-Delgado, 2008) and creating covariates Cellfraction_1, Cellfraction_2, Cellfraction_3, cellFrac_ilr_4. Transforming into the cell fractions has the additional advantage of invariance to scaling and to the changing the order of the variables.

### Differential Gene Expression Analysis

In order to gain insights into the pathophysiology associated with PTSD development, differential gene expression analysis was performed with the differential expression for repeated measures (dream) function available in the variancePartition R package (Hoffman and Schadt, 2020). Dream employs a linear model capable of increasing statistical power while minimizing false positives by joining multiple statistical concepts into a single statistical model implemented by its software (Hoffman and Roussos, 2020). The dream analysis is built onto the widely applied workflow for differential expression analysis of limma/voom (Law et al. 2014). The model includes several statistical concepts into its workflow including the modeling of repeated measures, precision weights capable of modeling measurement errors in the RNA-sequencing counts and the ability to include several random effects. Apart from the ability to model a range of random effects, there is a separate estimation of random effects for each single gene.

Next, the form for the differential gene expression analysis with the Dream function was specified in which the variable to be tested is required to be a fixed effect. For our analysis several metadata variables were included and assessed for their contribution to variation in differential expression. Those variables include disease status “PTSD”, Sex, age, time point (wave) as well as a range of cell proportions commonly considered in transcriptomic studies of whole blood samples from individuals with mental disorders including B cells, CD4T cells, CD8T cells and monocytes.

The first analysis focused on the cross sectional differences in gene expression of the PTSD affected group of participants vs those who are unaffected by PTSD. This analysis can yield insights into the transcriptomic differences between individuals affected by PTSD vs those who are not after the two timepoints. Here, the gene expression differences between cases and controls were tested first with and then without the inclusion of the wave covariate evaluate the potential contribution of the timepoint to the observed variation in differential expression. Subsequently, it was tested if the PTSD coefficient is different from zero in order to investigate if there is a PTSD effect indicating differential expression between cases and controls. For this cross-sectional analysis step the following regression formula was used first with and then without the “wave” covariate : ∼ ptsd + (1|resp) + wave + Sex + Age + Cellfraction_1 + Cellfraction_2 + Cellfraction_3 + Cellfraction_4. The “ptsd” term in this formula relates to the PTSD status at the end of wave 4, “resp” is the identifier for the individual participant, “wave” is the timepoint (w2= baseline, wave4= follow-up) and the cell fraction terms “Cellfraction_1 -Cellfraction_4” signify the log transformed cell proportion estimates.

Next, differential gene expression between cases and controls was assessed in a longitudinal analysis. The purpose of the longitudinal analysis was to identify differentially expressed transcriptomic signatures associated with the development of PTSD. In order to assess whether there are genes associated with the development of PTSD a wave-by-disease interaction was explored using the formula:

∼ ptsd + wave + ptsd:wave + (1|resp) + Sex +Age + Cellfraction_1 + Cellfraction_2 + Cellfraction_3 + Cellfraction_4

Subsequently, the coefficient PTSD:wave was tested for the association between PTSD and gene expression changes between the two waves by examining whether it was significantly different from 0 for each gene.

Additionally, differential gene expression was assessed cross-sectionally between the continuous PTS symptom severity measures of the participants instead of using the dichotomous PTSD variable.

This analysis intended to identify differentially expressed transcriptomic signatures in participants who experienced PTS but not to an extend that met the criteria for a PTSD diagnosis in line with DSM-IV. The analysis included the PCL score measuring PTS symptom severity with the following formula used first with and then without the “wave” covariate:

∼ (1|resp) + wave + Age + Sex + Cellfraction_1 + Cellfraction_2 + Cellfraction_3 + Cellfraction_4 + PCL_score_combined

Differential expression in our data was evaluated at a nominally significant p-value < 0.05 with a fold change of 1.5 or greater and also at an adjusted p-value < 0.05 using the Benjamini-Hochberg false discovery rate estimation (Hochberg and Benjamini, 1990).

### Gene Set Enrichment Analysis (GSEA)

In order to elucidate the biological processes underlying and accompanying PTSD development a gene set enrichment analysis (GSEA) was performed on the differentially expressed genes identified in the before and after the development of PTSD comparison. A GSEA can facilitate the identification of classes of genes found to be over-represented in a large gene set that can be subsequently associated with a disease. The R package ‘clusterProfiler’ can automate the process of biological-term classification and the enrichment analysis of gene clusters. We have applied the commonly used Gene Ontology (GO) (Ashburner et al., 2000) that provides annotations of genes to a range of biological processes, molecular functions, and different cell components. Differentially expressed signatures at a nominally significant p-value < 0.05 were tested for enrichment.

### Investigation of Differentially Expressed Signatures in the Human Brain

Many complex diseases underly the dysfunction of multiple tissues and organs. This dysfunction is often characterized by tissue specific transcriptional changes mediating causal links between the genotype and disease (Cookson et al., 2009). Assessing tissue specific gene expression is an important consideration given that the assessment of psychiatric disorders can be challenging because the organ central to the disease−the brain−is not available to be assessed in living individuals. Investigating if the differentially expressed genes identified in blood samples are also expressed in the brain can help evaluate how reflective blood transcriptomic profiles are of the disorders of the brain. To this end, we verified whether the differentially expressed genes found in blood are also expressed in the human brain by conducting a search on the publicly available Human Protein Atlas (HPA) database (http://www.proteinatlas.org/<u>).\</u>

## Results

### Study participants

Descriptive statistics for demographic and psychosocial measures in study participants are shown in Table 1 and with visual representations in Figure 1 and 2.

**Table 1.** Participant Characteristics of PTSD and Control Samples

|  | *All participants*<br>*(N = 72)* |  | *New Trauma exposure from*<br>*Baseline to follow-up*<br>*(N = 70)* |  | *PTSD-affected at Baseline*<br>*(N = 10)* |  | *PTSD-affected at follow-up (N = 31)* |  | *New Case of PTSD*<br>*(N = 21)* |  | *No PTSD by follow-up*<br>*(N = 41)* |  | *PTS symptom score at Baseline\*\**<br>*(Min-Max:6.0-72.0)* | *PTS symptom score by follow-up\*\**<br>*(Min-Max:17.0-73.0)* |
| --- | --- | --- | --- | --- | --- | --- | --- | --- | --- | --- | --- | --- | --- | --- |
| *Characteristic* | **N** | **%** | **N** | **%** | **N** | **%** | **N** | **%** | **N** | **%** | **N** | **%** |  |  |
| *Age\** | 56.66 | 12.78 | 56.27 | 12.55 | 56.98 | 12.80 | 56.49 | 12.94 | 54.38 | 13.91 | 56.97 | 12.10 |  |  |
| *Female* | 39 | 54 | 38 | 54 | 6 | 60 | 20 | 64 | 14 | 67 | 19 | 46 | 31.27 | 33.25 |
| *Male* | 33 | 46 | 32 | 46 | 4 | 40 | 11 | 36 | 7 | 33 | 22 | 54 | 32.46 | 33.83 |
| *African American* | 61 | 85 | 60 | 86 | 9 | 90 | 24 | 77 | 15 | 71 | 37 | 90 | 32.46 | 33.83 |
| *Caucasian* | 7 | 10 | 7 | 10 | 1 | 10 | 5 | 16 | 4 | 19 | 2 | 5 | 31.16 | 31.13 |
| *Other* | 4 | 5 | 3 | 4 | 0 | 0 | 2 | 7 | 2 | 10 | 2 | 5 | 30.66 | 32.21 |

\*Variables are presented by mean and standard deviation. \*\*Variables are presented by mean.

**Figure 1.**
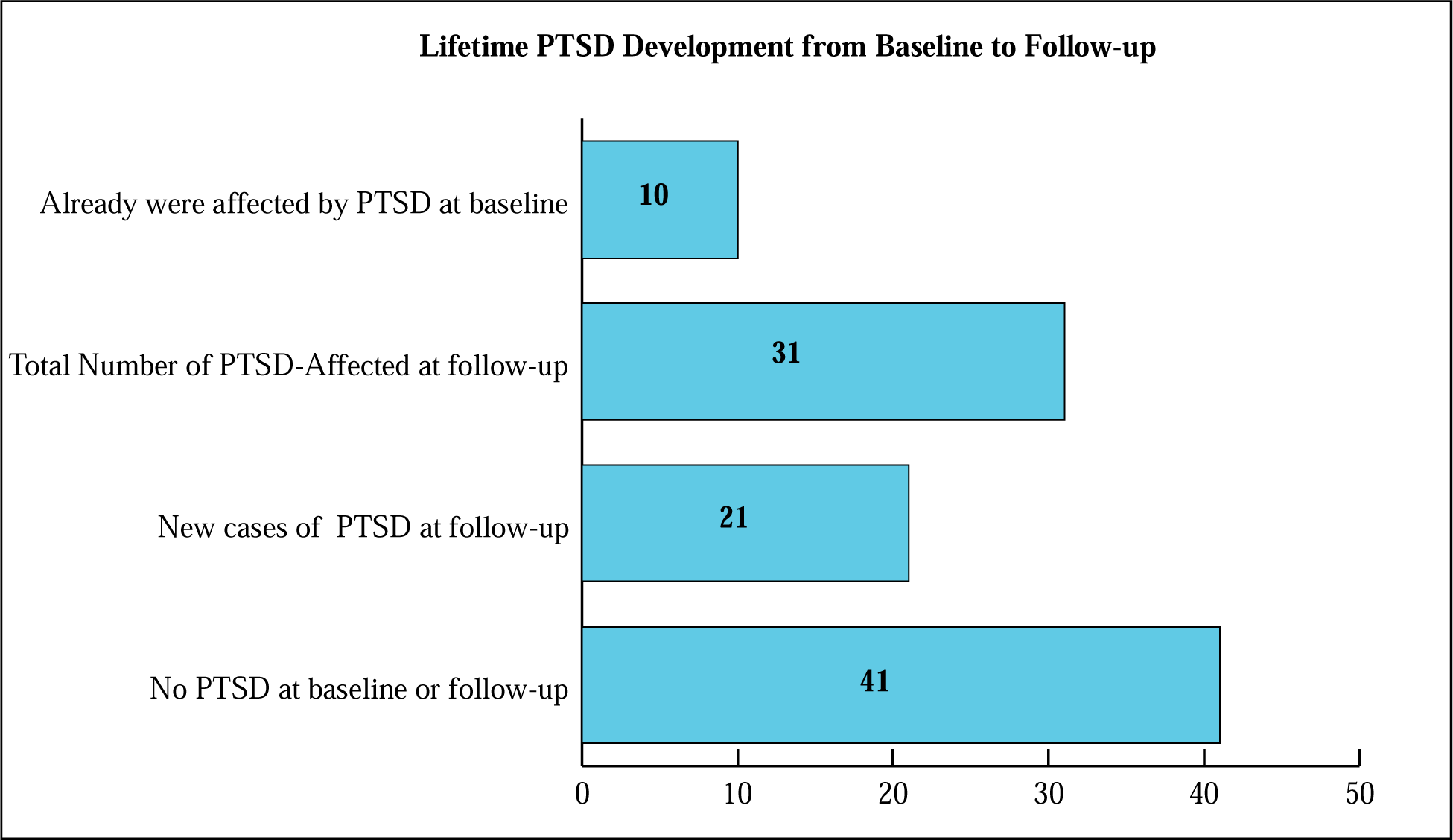
Breakdown of number of participants with PTSD

**Figure 2.**
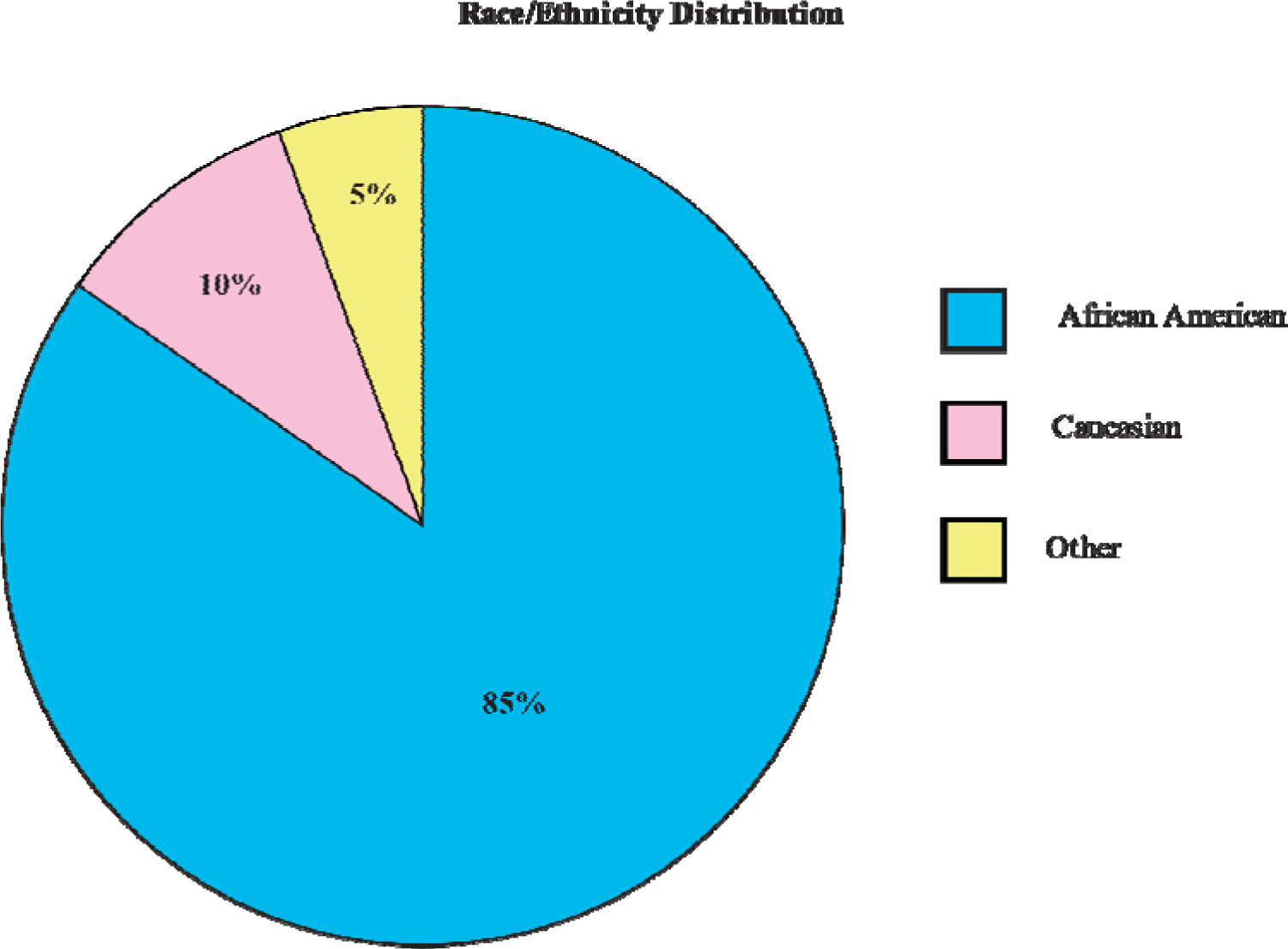
Demographic breakdown of study participants

### Outcome of RNA processing, gene mapping and gene assignment

From the original RNAseq experiment, a total of 320,197,33 reads were obtained mapping to 58,884 unique transcripts. 75% of obtained reads successfully mapped to the GRCh38 reference genome (ftp://ftp.ebi.ac.uk/pub/databases/gencode/Gencode_human/release_24/GRCh38.primary_assembly.gen ome.fa.gz). 87% of reads were assigned to a total of 26,659 individual genes after mapping with the use of the FeatureCounts function of the R-package Subreads (Liao, Smyth & Shi, 2019).

### Estimation of the Blood Cell Type Proportions

Using quality-controlled data, Cibersortx was used to determine the cell-type composition of the heterogenous blood samples including the baseline (Figure 3) and the follow-up (Figure 4) transcriptomic measures. For both timepoints the quantified proportions included B cells, CD4T, CD8T, Monocytes and NK cells for each participant. Natural killer T cell estimates were provided in the Cibersortx output but not included in the exploration of covariates among the cell proportions because they were only represented in less than 4% of the samples.

**Figure 3.**
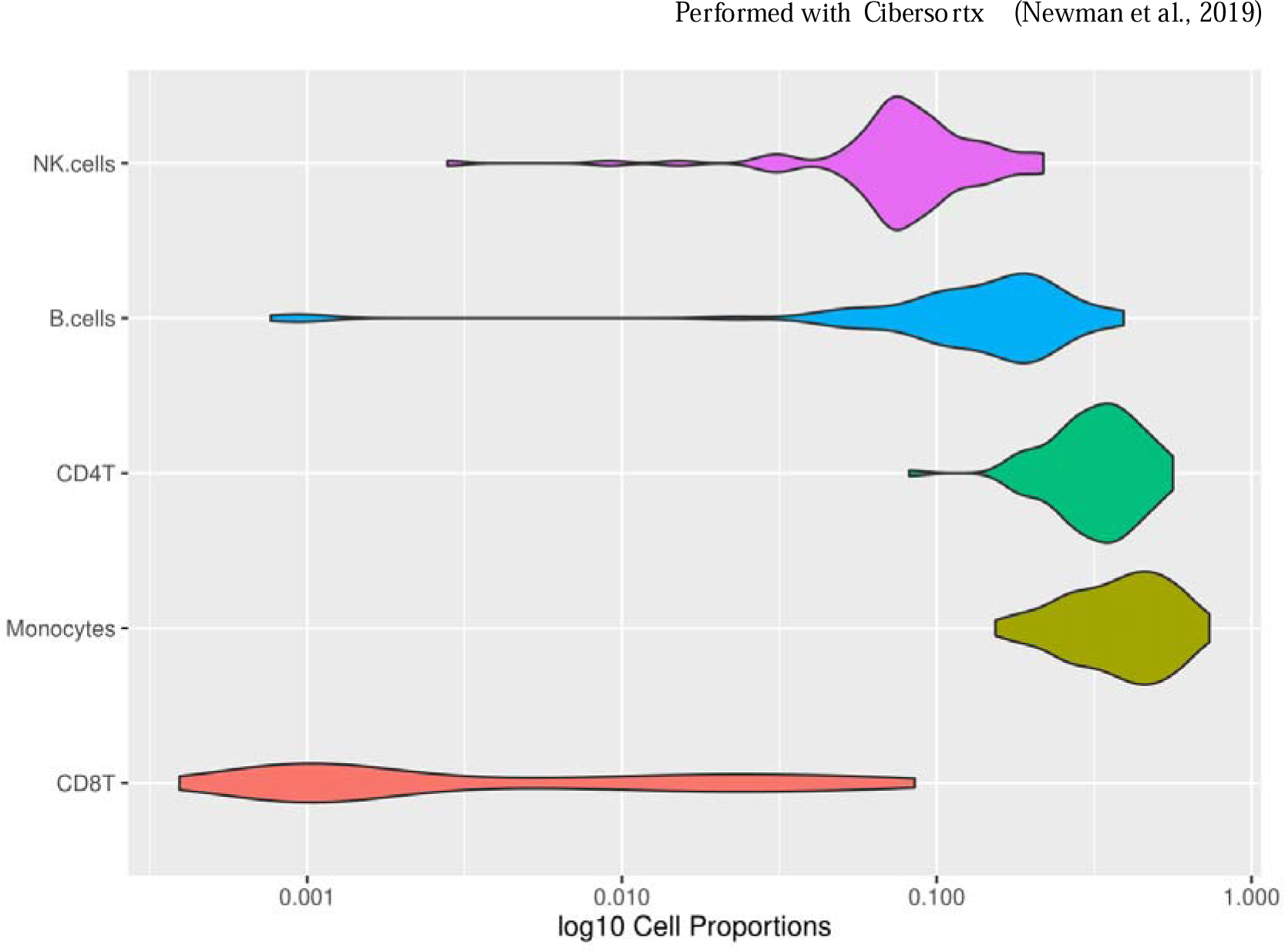
Cell proportions at the baseline measure Cell proportions of the 72 samples included in the study at the baseline measure including Cell fractions of B cells, CD4T, CD8T, Monocytes, NK cells.

**Figure 4.**
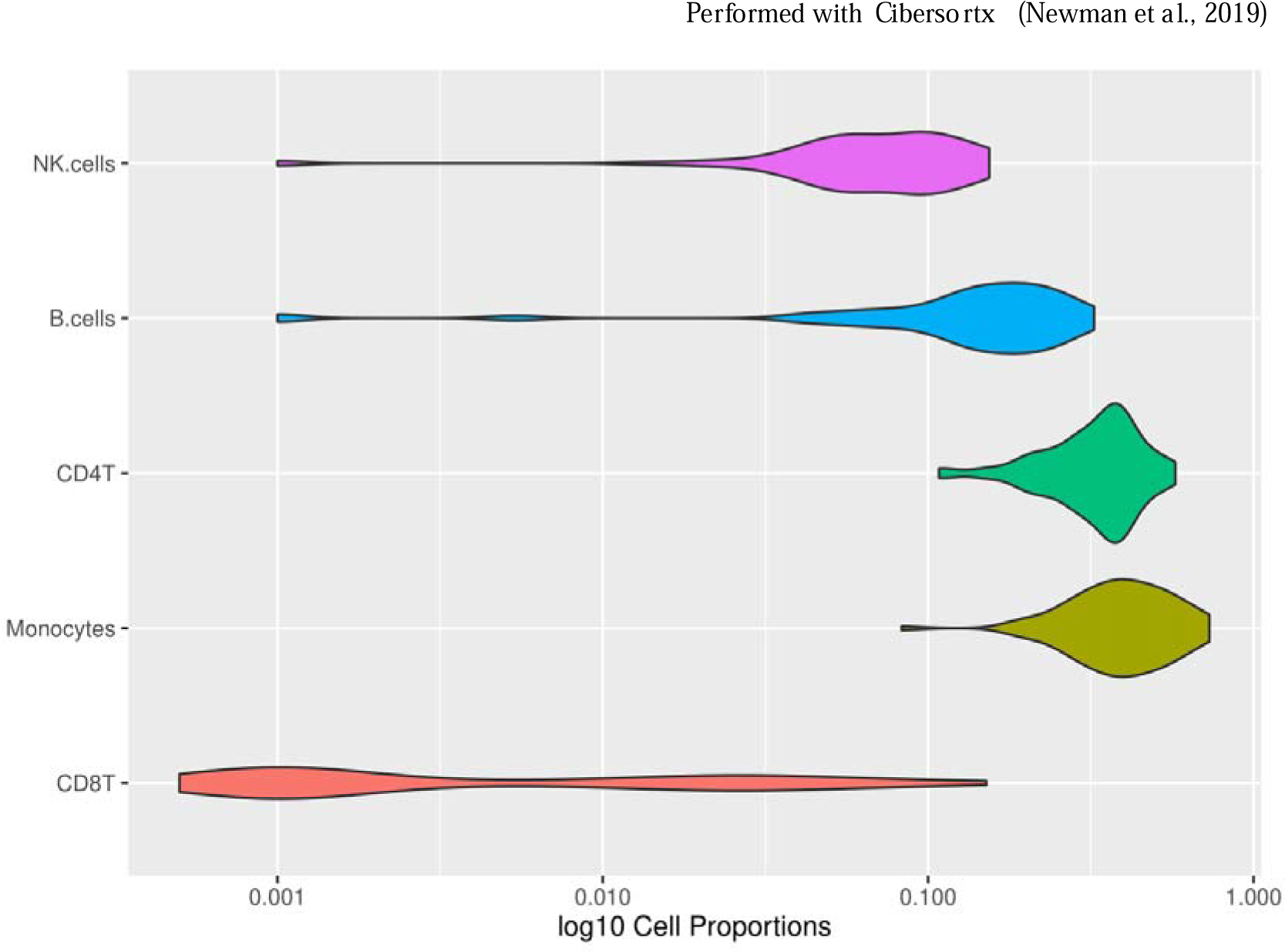
Cell fractions of the baseline measure displayed in a violin plot

### Differential Gene Expression

To gain insight into the gene expression signatures associated with PTSD development in our community-based cohort, a differential gene expression analysis was performed testing multiple coefficients of interest. Retention of power and the controlling of the false positivity rate was ensured by accounting for the repeated measures of our PTSD cases and controls with the dream function of the variancePartition R package (Hoffman et al., 2020), which enables the specification of random effects. In the dream analysis, the effects of the individual on the gene expression are modeled as a random effect for each individual gene with the use of a linear mixed model. Different differential gene expression analyses were performed to investigate the transcriptomic differences between participants affected by PTSD vs those who are not as well as to identify differential gene expression associated with the development of PTSD and PTS symptom severity. For the models, multiple test correction was performed taking the number of genes tested into account with the use of the Benjamini and Hochberg method (Hochberg and Benjamini, 1990).

First, the gene expression between cases vs controls was tested with and without the inclusion of the wave covariate in order to examine the cross-sectional gene expression differences in individuals who developed PTSD vs those who did not. Subsequently, it was tested if the PTSD coefficient is different from zero in order to investigate if there is a PTSD effect. Neither of these analyses identified any differentially expressed genes with a log^2^fold change greater than 1.5 at an unadjusted p-value < 0.05. Similarly, testing for gene expression that included the PCL score measuring PTS symptom severity instead of PTSD, did not identify any differentially expressed genes even at the nominal p-value < 0.05.

In contrast, the analysis testing for a wave-by-disease interaction and subsequently testing the coefficient “PTSD:wave” for the association between PTSD and gene expression changes between the two waves identified several differentially expressed genes with a log2fold change greater than 1.5 at an unadjusted p-value < 0.05. The volcano plot (Figure 5) shows the global gene expression patterns according to PTSD development, indicating a larger number of upregulated genes compared to a few downregulated genes associated with PTSD development. In total, 45 genes were identified as differentially expressed at a nominal p-value < 0.05 in relation to PTSD development. None of the differentially expressed genes survived multiple test correction at an adjusted p-value < 0.05.

**Figure 5.**
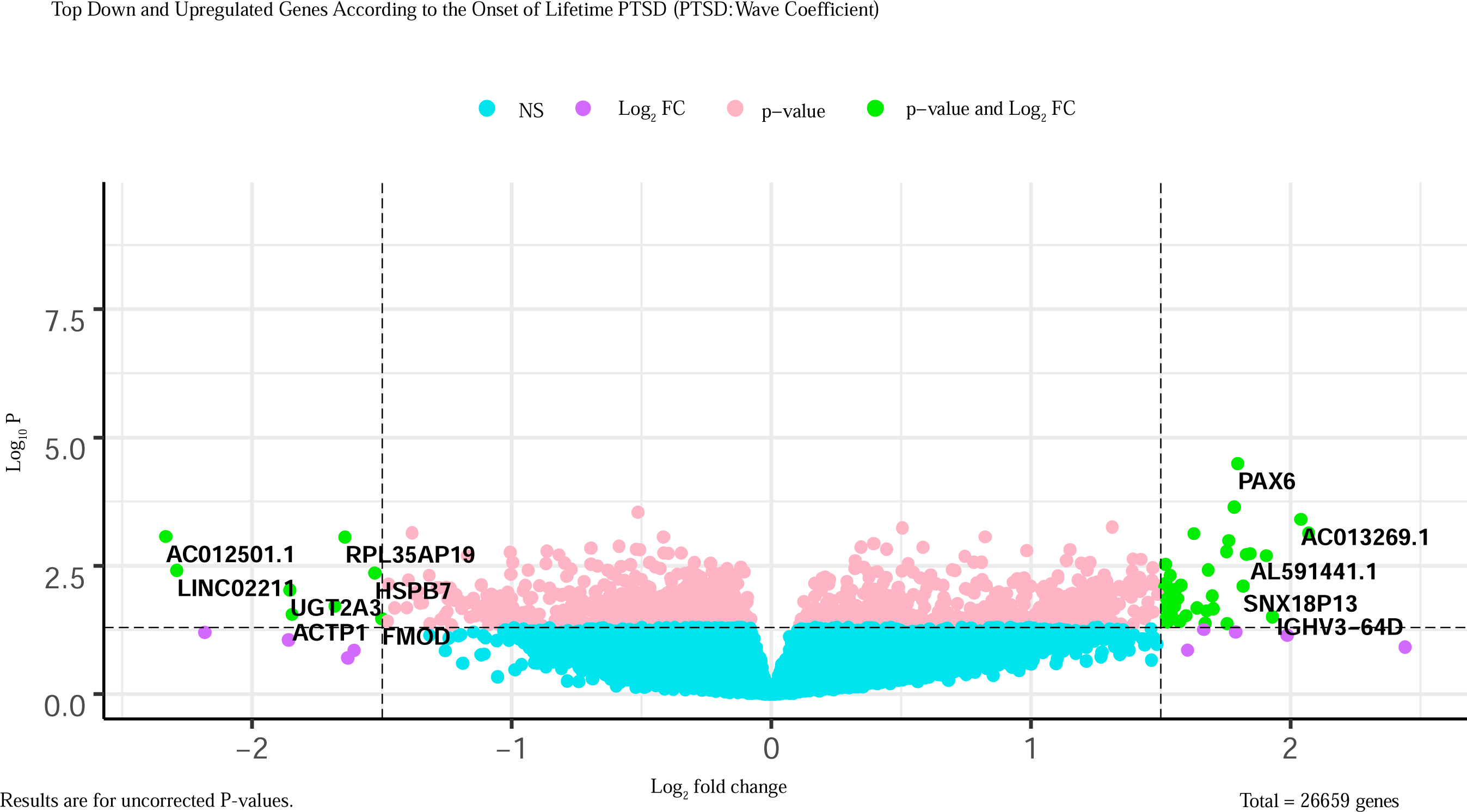
Volcano plot displaying differentially expressed genes according to Lifetime PTSD Development

There were a total of 37 upregulated genes associated with PTSD development at a nominal p-value < 0.05 and log_2_ (fold change) above 1.5 (Table 2). Among the top differentially expressed genes with the highest fold change were *AL136317.2*, *AC013269.1*, *IGHV3-64D*, *AF130359.1*, *AL591441.1*, *TSPAN7*, *SNX18P13*, *PAX6*, *AC008543.5*, *Z69706.1* and *PXDN*. Furthermore, eight genes including *AC012501.1*, *LINC02211*, *UGT2A3*, *ACTP1*, *LINC02228*, *RPL35AP19*, *HSPB7* and *FMOD* were identified as downregulated according to PTSD development (Table 3).

**Table 2.** Upregulated Genes According to Lifetime PTSD Development

*Upregulated at Nominal p-value < 0.05 in PTSD:wave (log fold change 1.5)*
| *Ensemblid* | *Gene* | *LogFC* | *AveExpr* | *t* | *P.Value* | *Adj.P.Val* | *Z.std* |
| --- | --- | --- | --- | --- | --- | --- | --- |
| *ENSG00000274893* | *AL136317.2* | 2.069513 | -3.80093 | 3.476319 | 0.00073 | 0.952575 | 3.37795 |
| *ENSG00000225341* | *AC013269.1* | 2.039292 | -4.14782 | 3.665617 | 0.000393 | 0.952575 | 3.54454 |
| *ENSG00000282639* | *IGHV3-64D* | 1.929707 | -3.0084 | 2.187479 | 0.031035 | 0.952575 | 2.156621 |
| *ENSG00000237735* | *AF130359.1* | 1.906208 | -4.58473 | 3.175578 | 0.002008 | 0.952575 | 3.089076 |
| *ENSG00000276128* | *AL591441.1* | 1.844134 | -4.40663 | 3.198495 | 0.00182 | 0.952575 | 3.118174 |
| *ENSG00000156298* | *TSPAN7* | 1.82917 | -3.79308 | 3.170037 | 0.001889 | 0.952575 | 3.107136 |
| *ENSG00000230965* | *SNX18P13* | 1.816912 | -2.16646 | 2.708481 | 0.007875 | 0.952575 | 2.657394 |
| *ENSG00000007372* | *PAX6* | 1.796315 | -3.00469 | 4.301555 | 3.24E-05 | 0.877348 | 4.155555 |
| *ENSG00000267646* | *AC008543.5* | 1.782542 | -3.37977 | 3.818061 | 0.000227 | 0.952575 | 3.686855 |
| *ENSG00000268836* | *Z69706.1* | 1.761723 | -3.34508 | 3.357925 | 0.001023 | 0.952575 | 3.284242 |
| *ENSG00000130508* | *PXDN* | 1.754969 | -1.95644 | 2.052409 | 0.042077 | 0.952575 | 2.032763 |
| *ENSG00000285079* | *AL513493.1* | 1.752864 | -3.2731 | 3.208111 | 0.001672 | 0.952575 | 3.143115 |
| *ENSG00000184761* | *PRSS40B* | 1.702262 | -3.44557 | 2.328637 | 0.021705 | 0.952575 | 2.295487 |
| *ENSG00000187135* | *VSTM2B* | 1.698038 | -3.62639 | 2.540431 | 0.012212 | 0.952575 | 2.505954 |
| *ENSG00000229847* | *EMX2OS* | 1.681767 | -1.7885 | 2.961896 | 0.003813 | 0.952575 | 2.893211 |
| *ENSG00000185736* | *ADARB2* | 1.676024 | 0.873709 | 2.288869 | 0.023651 | 0.952575 | 2.262747 |
| *ENSG00000261348* | *LINC02136* | 1.670426 | -4.42673 | 2.06117 | 0.04176 | 0.952575 | 2.035902 |
| *ENSG00000170153* | *RNF150* | 1.639384 | -1.0087 | 2.346386 | 0.020885 | 0.952575 | 2.310054 |
| *ENSG00000226676* | *BX248123.1* | 1.627546 | -3.69832 | 3.487307 | 0.000746 | 0.952575 | 3.372131 |
| *ENSG00000235726* | *AC010148.1* | 1.597349 | -1.52826 | 2.203848 | 0.029654 | 0.952575 | 2.174677 |
| *ENSG00000286099* | *N/A* | 1.592427 | -2.53084 | 2.166485 | 0.032043 | 0.952575 | 2.143873 |
| *ENSG00000188730* | *VWC2* | 1.578 | -4.36536 | 2.713234 | 0.007538 | 0.952575 | 2.672072 |
| *ENSG00000145147* | *SLIT2* | 1.571715 | -0.69046 | 2.099633 | 0.03817 | 0.952575 | 2.07303 |
| *ENSG00000228314* | *CYP4F29P* | 1.563679 | 0.328895 | 2.503854 | 0.013714 | 0.952575 | 2.464657 |
| *ENSG00000233048* | *LINC01722* | 1.555737 | -3.63935 | 2.646673 | 0.009102 | 0.952575 | 2.608181 |
| *ENSG00000137573* | *SULF1* | 1.553857 | -4.42013 | 2.379621 | 0.019074 | 0.952575 | 2.344088 |
| *ENSG00000228971* | *AL356479.1* | 1.540334 | -4.04196 | 2.219536 | 0.028132 | 0.952575 | 2.195446 |
| *ENSG00000228791* | *THRB-AS1* | 1.53563 | -3.00011 | 2.878841 | 0.004867 | 0.952575 | 2.815727 |
| ENSG00000225258 | AC009478.1 | 1.529383 | -4.83787 | 2.666998 | 0.008854 | 0.952575 | 2.615629 |
| ENSG00000232006 | AC005537.1 | 1.525101 | -2.33867 | 2.087177 | 0.03932 | 0.952575 | 2.060819 |
| ENSG00000276484 | AC126755.5 | 1.524739 | -3.30051 | 2.088198 | 0.038673 | 0.952575 | 2.067652 |
| ENSG00000227766 | HCG4P5 | 1.522675 | -5.30751 | 2.497207 | 0.013944 | 0.952575 | 2.45871 |
| ENSG00000060718 | COL11A1 | 1.521923 | -4.74859 | 2.361928 | 0.020029 | 0.952575 | 2.325796 |
| ENSG00000135903 | PAX3 | 1.521438 | -4.30189 | 2.17104 | 0.032099 | 0.952575 | 2.143173 |
| ENSG00000100968 | NFATC4 | 1.519876 | -3.85836 | 2.744275 | 0.006896 | 0.952575 | 2.701825 |
| ENSG00000213614 | HEXA | 1.517953 | -4.52251 | 3.040019 | 0.002968 | 0.952575 | 2.97099 |
| ENSG00000168122 | ZNF355P | 1.515112 | -4.04418 | 2.693571 | 0.008234 | 0.952575 | 2.642316 |

**Table 3.** Downregulated Genes According to Lifetime PTSD Development

*Downregulated at Nominal p-value < 0.05 in PTSD:wave (log fold change 1.5)*
| Ensemblid | Gene | LogFC | AveExpr | t | P.Value | Adj.P.Val | Z.std |
| --- | --- | --- | --- | --- | --- | --- | --- |
| ENSG00000227400 | AC012501.1 | -2.33236 | -2.16611 | -3.41908 | 0.000845 | 0.952575 | -3.33756 |
| ENSG00000245662 | LINC02211 | -2.28962 | -3.46875 | -2.94785 | 0.00389 | 0.952575 | -2.88695 |
| ENSG00000135220 | UGT2A3 | -1.85419 | -4.07665 | -2.64266 | 0.009397 | 0.952575 | -2.59725 |
| ENSG00000229890 | ACTP1 | -1.84626 | -4.4253 | -2.22633 | 0.027948 | 0.952575 | -2.19802 |
| ENSG00000251273 | LINC02228 | -1.68067 | -4.02054 | -2.37971 | 0.019097 | 0.952575 | -2.34364 |
| ENSG00000239872 | RPL35AP19 | -1.64212 | -4.78866 | -3.41661 | 0.000874 | 0.952575 | -3.32825 |
| ENSG00000173641 | HSPB7 | -1.52756 | -4.55159 | -2.90137 | 0.004345 | 0.952575 | -2.85199 |
| ENSG00000122176 | FMOD | -1.50034 | -4.45216 | -2.15001 | 0.033614 | 0.952575 | -2.12468 |

**Table 4.** Differentially expressed Genes expressed in the Brain

| Gene | Log2fc | Expression in human Brain | Implicated in previous Studies of psychiatric disorders |
| --- | --- | --- | --- |
| *TSPAN7* | 1.82917 | All regions of the brain | Not implicated in psychiatric disorders |
| *PAX6* | 1.796315 | All regions of the brain | Important transcriptional factor for normal brain development and is involved in the regulation of the expression of several other genes coordinating cortical development (Osumi et al., 2008) |
| *PXDN* | 1.754969 | All regions of the brain | Differentially methylated CpG in PXDN associated with borderline personality disorder (Arranz et al., 2021) |
| *VWC2* | 1.578 | High expression cerebellum | Differentially expressed (downregulated) in individuals with PTSD (Kuan et al., 2020). |
| *SULF1* | 1.553857 | High expression in thalamus and midbrain | Differential methylation of SULF1 associated with depression during pregnancy (Viuff et al., 2018) |
| *PAX3* | 1.521438 | High expression in cerebellum and white matter | Not implicated in psychiatric disorders |
| *NFATC4* | 1.519876 | High expression in cerebellum and white matter | Dysregulation of other *NFATC* loci has previously been implicated in PTSD in peripheral tissues (Breen et al. 2019; Occean et al. 2022) |
| *HSPB7* | -1.52756 | Higher expression in medulla oblongata, pons and cerebellum | Previously been associated with the development of Schizophrenia (Chaumette et al., 2019) |
| *FMOD* | -1.50034 | Higher expression in cerebral cortex | Not implicated in psychiatric disorders |

### Gene Set Enrichment Analysis

In order to identify the biological processes associated with PTSD development, a gene set enrichment analysis was performed. Investigating the pathway enrichment for the 37 upregulated and eight downregulated differential expression signatures at the nominal significant p-value < 0.05 we found some gene sets to be significantly enriched. The top gene ontology terms associated with PTSD development in regard to the PTSD:wave measure (Figure 6 and Figure 7) were found to be involved in cellular organization and functioning.

**Figure 6.**
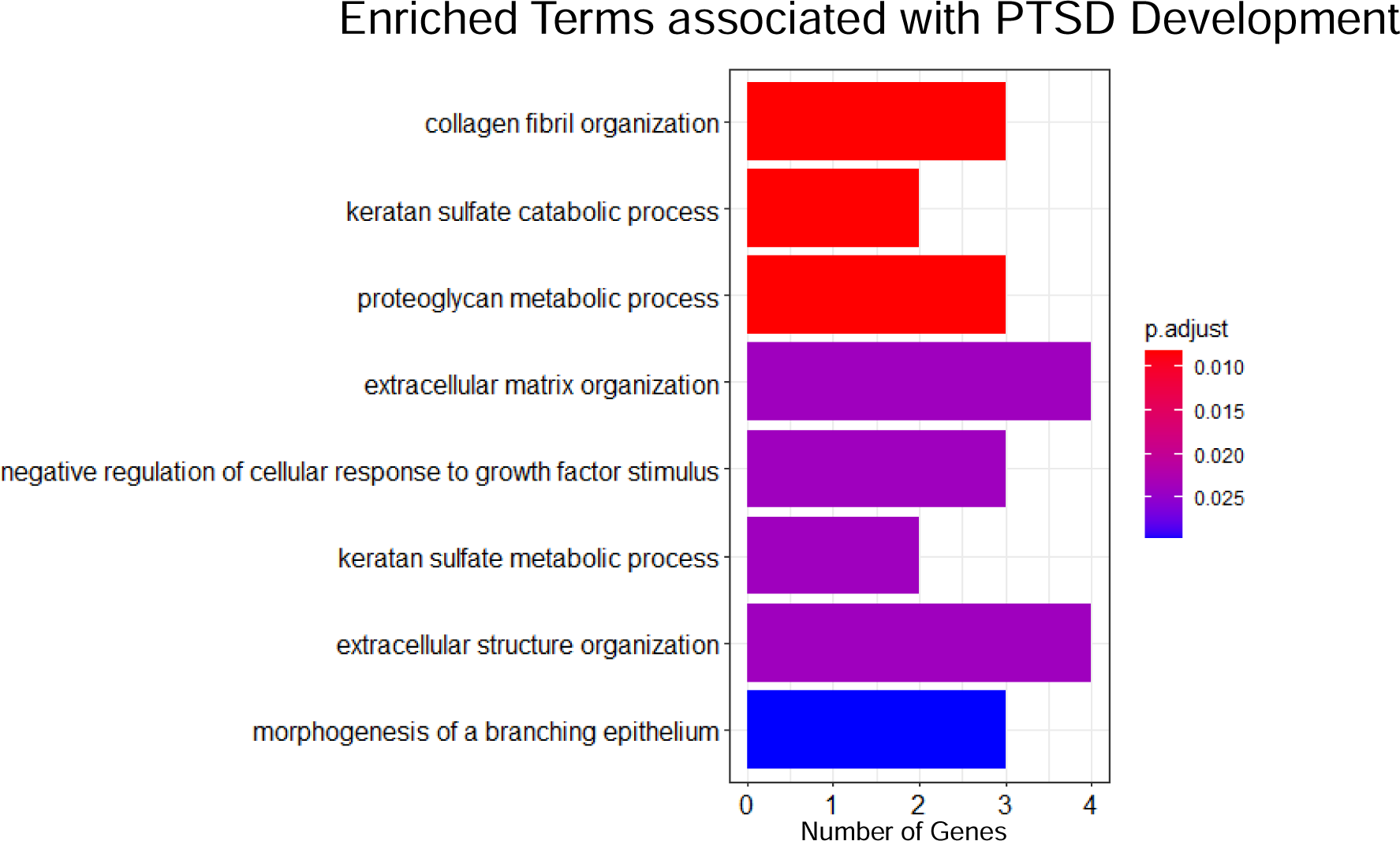
Top gene ontology terms associated with PTSD development

**Figure 7.**
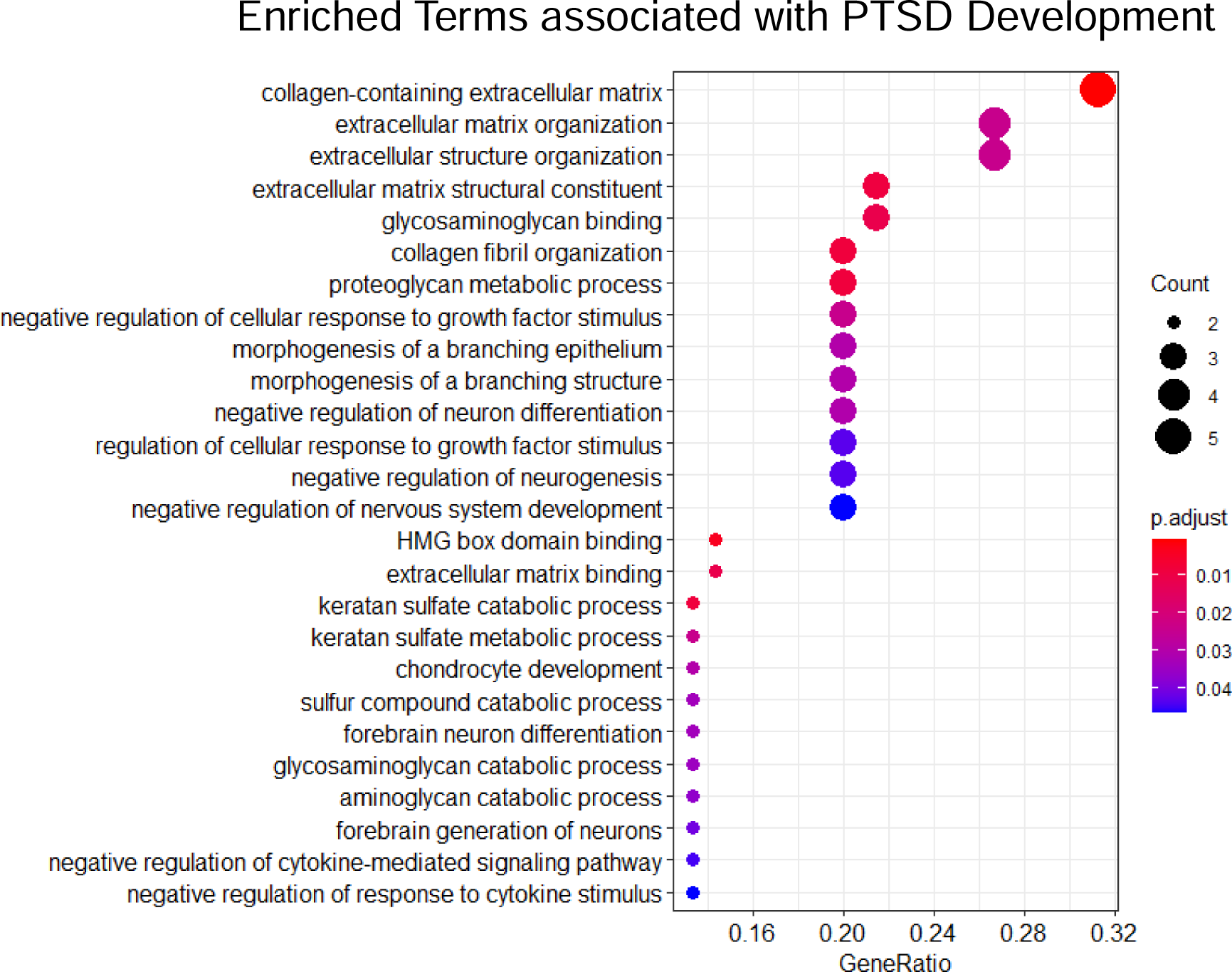
Dotplot of gene ontology terms in relation to PTSD development

### Confirmation of blood-based differentially expressed signatures in the human brain

We leveraged the protein atlas database to assess whether our DE genes identified in blood are also expressed in the brain, a key organ of interest in PTSD that cannot ordinarily be accessed in living individuals. Several of our differentially expressed genes identified in blood are also expressed in the brain (Table 3). With regard to the upregulated genes, *TSPAN7, PAX6, PXDN, VWC2, SULF1, PAX3* and *NFATC4* were also expressed in the brain. Out of the downregulated genes, *UGT2A3*, *ACTP1*, *HSPB7* and *FMOD* were also found to be expressed in the brain.

## Discussion

Despite the contributions of several RNA-sequencing studies of PTSD, longitudinal analysis that characterizes the gene expression changes associated with the development of the disorder are lacking. In an attempt to address this gap in knowledge, we analyzed RNA-seq data in whole blood from 72 community-dwelling individuals (males and females) from the Detroit Neighborhood Health Study. Among them, 21 developed PTSD from the baseline to the follow-up time point. A linear mixed model optimized for repeated measures was applied to analyze the gene expression data. We found 45 differentially expressed genes in our cohort of individuals who developed PTSD with an estimated log_2_fold change > 1.5 at a nominal p value; however, none of these survived correction for multiple hypothesis testing. Gene set enrichment analysis of the differentially expressed genes implicated several pathways to be enriched in individuals developing PTSD. The top enriched pathways included brain functions such as forebrain neuron differentiation and the regulation of neurogenesis. Additionally, two of the top enriched pathways were involved with the negative regulation of cytokine-mediated signaling and the negative regulation of the response to cytokine stimulus, immune-related pathways that have been implicated in PTSD in previous studies (Breen et al., 2015; Bam et al., 2016; Kuan et al., 2020). PTSD has been consistently linked to increased inflammation in the body, which is in line with the findings of some of our immune-related enriched pathways (Michopolous et al., 2017; Rosen et al., 2017).

Only one previous study by Breen and colleagues (Breen et al., 2015) complemented their cross-sectional gene expression analysis with the longitudinal assessment of the transcriptome in a pre- and post-deployment setting of male U.S Marines. Although the majority of the observed changes in gene expression occurred cross-sectionally between the timepoints pre- and post-deployment, the authors observed 57 upregulated and 53 downregulated genes in their longitudinal analysis at nominally (p-value < 0.05) significant thresholds. None of the identified differentially expressed genes from their longitudinal contrast analysis between pre- and post-deployment survived multiple test correction at an adjusted p-value < 0.05 but were nonetheless included in a subsequent network analysis. Their network analysis revealed dysregulated modules relating to functions of the innate immunity associated with PTSD development. Those differences observed in the innate immunity modules were not merely observed cross sectionally as a consequence of PTSD but also of putative causal significance for the development of the disorder. Despite their comprehensive study design, most of the differentially expressed genes identified in their analyses that were basis for their network analysis were observed cross sectionally, limiting insights into transcriptomic mechanisms associated with the development of the disorder. Further, their cohort of U.S marines was limited to Caucasian males leaving room for studies that include females and other ethnicities.

Assessing transcriptional alterations directly in the brain is not possible in living patients with PTSD and stimulates the broader question of to which extent gene expression signatures observed in peripheral tissue reflect the organ central to PTSD-the brain. To this end, we assessed whether our DE genes identified in blood are also expressed in the brain. Several of our differentially expressed genes identified in blood are also expressed in the brain (Human Protein Atlas (HPA) database (http://www.proteinatlas.org/) With regard to the upregulated genes, *TSPAN7, PAX6, PXDN, VWC2, SULF1, PAX3* and *NFATC4* were also expressed in various regions of the brain. While *TSPAN7, PAX6, PXDN* were ubiquitously expressed in all regions of the brain, *PAX3* and *NFATC4* showed higher expression in the cerebellum and in the white matter of the brain. *VWC2* on the other hand, has its highest expression in the cerebellum and only low expression in other areas of the brain. Out of the downregulated genes, *HSPB7* and *FMOD* were also found to be expressed in the brain. While *HSPB7* showed its highest expression in the medulla oblongata, pons and the cerebellum, *FMOD* appeared to have more expression in the cerebral cortex. *PAX6* is an important transcriptional factor for normal brain development and is involved in the regulation of the expression of several other genes coordinating cortical development (Osumi et al., 2008; Ypsilanti & Rubenstein, 2016). *TSPAN7* has been found to be differentially expressed in a previous microarray study of peripheral blood investigating the gene expression profiles of military personnel who sustained closed-head injuries from explosions during deployment (Heinzelmann et al., 2014). *TSPAN7* has also been associated with cognitive defects mediated by downstream alterations to the *TM4SF2* gene (Penzes et al., 2013). *VWC2* has been previously found to be differentially expressed (downregulated) in individuals with PTSD and mild cognitive impairment in a targeted study of World Trade Center responders (Kuan et al., 2020). *NFATC4* is among a family of five distinct *nuclear factor of activated T cells* genes regulating the transcriptional induction of several genes involved with immune activators and modulators including a range of adaptive immunity and pro-inflammatory cytokines (Kipanyula, Kimaro & Etet, 2016). Dysregulation of other *NFATC* loci has previously been implicated in PTSD in peripheral tissues (Breen et al. 2019). Although there was no direct overlap of the identified differentially expressed genes in our community-dwelling cohort and the U.S marine cohort investigated by Breen and Colleagues, our results confirm the dysregulation of genes and the enrichment of pathways involved with immune functioning previously implicated in other PTSD studies.

No differentially expressed genes were observed after multiple test correction for any of the models we tested. This outcome is not surprising considering the efforts to prevent false positives by accounting for several covariates and our small sample size. Although no statistically significant results were obtained in our cohort, the longitudinal sampling of participants prior to developing PTSD can be important in identifying expression signatures underlying PTSD development in future studies. A strength of our study is the assessment of a community-based cohort as opposed to accessing PTSD in veterans, active-duty military or first responders. Additionally, this work might be of use in larger meta-analysis efforts that leverage larger sample sizes to identify transcriptomic signatures underlying or accompanying the development of PTSD. Further, the implicated genes associated with PTSD development could be evaluated in future genome wide association studies. With regard to embodiment, the longitudinal assessment of transcriptomic signatures associated with PTSD development can help elucidate the complex process of how traumatic experiences are embodied into our physiology and ultimately reflected in psychopathology.

## Data Availability

All data produced in the present study are available upon reasonable request to the authors.

https://github.com/dahrendorff/Longitudinal_RNAseq_PTSD/blob/main/Longitudinal_RNAseq_PTSD.R

## Acknowledgements

We are grateful to the study staff and volunteers of the Detroit Neighborhood Health Study for their contributions and appreciate the many Detroit residents who participated in the DNHS.

This study was supported by the National Institutes of Health Grants [R01DA022720, R01DA022720-S1, RC1MH088283, R01 MD011728]. RNA sequencing was supported by the Illinois Health Disparities Institute and performed by the DNA Services Lab at the University of Illinois at Urbana-Champaign. J. Dahrendorff was supported by the COPH Doctoral Fellowship of the University of South Florida.

